# VentRa. Validation study of the ventricle feature estimation and classification tool to differentiate behavioral variant frontotemporal dementia from psychiatric disorders and other degenerative diseases

**DOI:** 10.1101/2022.03.29.22273096

**Authors:** Ana L. Manera, Mahsa Dadar, Simon Ducharme, D. Louis Collins

## Abstract

**Introduction:** Lateral ventricles are reliable and sensitive indicators of brain atrophy and disease progression in behavioral variant frontotemporal dementia (bvFTD). Here we validate our previously developed automated tool using ventricular features (known as VentRa) for the classification of bvFTD vs a mixed cohort of neurodegenerative, vascular, and psychiatric disorders from a clinically representative independent dataset.

**Methods:** Lateral ventricles were segmented for 1110 subjects - 14 bvFTD, 30 other Frontotemporal Dementia (FTD), 70 Lewy Body Disease (LBD), 898 Alzheimer Disease (AD), 62 Vascular Brain Injury (VBI) and 36 Primary Psychiatric Disorder (PPD) from the publicly accessible National Alzheimer’s Coordinating Center dataset to assess the performance of VentRa.

**Results:** Using ventricular features to discriminate bvFTD subjects from PPD, VentRa achieved an accuracy of 84%, 71% sensitivity and 89% specificity. VentRa was able to identify bvFTD from a mixed age-matched cohort (i.e., Other FTD, LBD, AD, VBI and PPD) and to correctly classify other disorders as ‘not compatible with bvFTD’ with a specificity of 83%. The specificity against each of the other individual cohorts were 80% for other FTD, 83% for LBD, 83% for AD and 84% for VBI.

**Discussion:** VentRa is a robust and generalizable tool with potential usefulness for improving the diagnostic certainty of bvFTD, particularly for the differential diagnosis with PPD.

## 1. INTRODUCTION

A confirmed behavioural variant frontotemporal dementia (bvFTD) diagnosis is often difficult to achieve in the clinic and heavily relies on brain imaging. This is explained in part by the high heterogeneity in heritability, pathology, and the absence of a unique molecular biomarker. In addition, the symptomatic overlap with other neurodegenerative disorders, but more importantly with non-degenerative primary psychiatric disorders (PPD) - including major depressive disorder, bipolar disorder, schizophrenia, obsessive-compulsive disorder, autism spectrum disorders and even personality disorders^1^ - means that PPD often constitutes the main differential diagnosis of bvFTD^2^. It has been reported that approximately 50% of bvFTD patients receive a prior psychiatric diagnosis (most frequently major depression), and the average diagnostic delay is up to 5-6 years from symptom onset ^1 3^. It has also been shown that patients with PPD can be wrongly diagnosed with bvFTD, particularly in community settings, preventing patients to have access to evidence-based psychiatric treatments^4^.

In previous work, using deformation-based morphometry (DBM), we showed that the ventricles are a robust metric to discriminate bvFTD from cognitively normal controls (CN)^5^. The lateral ventricles exhibited the most significant volumetric difference at baseline between bvFTD and CN and, of all structures, they showed the most significant progression of change in 1-year follow up.

In more recent work, we assessed the differences in ventricular features between bvFTD, normal controls, and other dementia cohorts and developed VentRa, the first tool to use ventricular features specifically for the differential diagnosis of bvFTD. These features included total ventricular volume (TVV), left-right frontal ventricle volume ratio (LRFR), left-right temporal ventricle volume ratio (LRTR) and the anterior-posterior ratio (APR). Using VentRa, we achieved a 10-fold cross-validation accuracy of 80% (76% sensitivity, 83% specificity) in differentiating bvFTD from all other cohorts that included normal aging, mild cognitive impairment (MCI), Alzheimer’s dementia (AD), and other Frontotemporal Dementia (FTD) variants. Interestingly, using only the APR ventricular feature yields a 76% accuracy, underlying its importance in differentiating between groups^6^. The lateral ventricles are easy to reliably segment manually or when using standard publicly available tools ^7 8^ and provide an estimate of the overall extent of brain atrophy across different regions, making the VentRa ventricle-based features for bvFTD diagnosis a promising practical tool both in research and clinical settings.

In the present study, we aimed to validate VentRa on an independent cohort obtained from the National Alzheimer’s Coordinating Center dataset (NACC) to differentiate bvFTD subjects from other FTD, Lewy Body Disease (LBD), AD, Vascular Brain Injury (VBI) and PPD. While the key differential diagnostic is between bvFTD variants and PPD, we assessed the performance of VentRa in data from patients with other neurodegenerative dementias (other FTD subtypes, AD, LBD, and VBD) to verify that VentRa can detect atrophy patterns specific to bvFTD and not just any neurodegenerative disease related pattern of atrophy that might also occur in other dementias. The publicly available NACC dataset was selected as the target dataset for independent validation of VentRa since it contains a large number of multi-center and multi-scanner datasets with T1-weighted images acquired with different acquisition protocols, field of strength, resolution, and field of view, allowing us to validate the generalizability of VentRa in realistic clinical settings.

## 2. MATERIALS AND METHODS

### 2.1 Participants

The data sample used for this study was obtained from the National Alzheimer’s Coordinating Center’s Uniform Data Set (NACC). The NACC developed a database of standardized clinical research data obtained from past and present National Institute on Aging (NIA)-funded Alzheimer’s Disease Research Centers across the United States. NACC data collection has been previously described elsewhere^9^. Participants included in the database have a range of cognitive statuses: normal cognition, MCI, and demented due to various clinical diagnoses. A total of 5246 scans were obtained from NACC from the September 2019 data freeze after a data request for all participants with any dementia syndrome (i.e., bvFTD, AD, VBI, LDB and other), MCI and cognitively normal subjects who have at least one T1 weighted (T1-W) scan ± 6 months from a clinical visit [NACC variables MRIT1=1 and NACCMRDY between (−180 and 180)]. Figure 1 shows the flowchart for the detailed subject selection procedure. The analysis reported here used data from 1110 subjects from the six diagnostic groups of interest – bvFTD (n=14), other FTD (n=30), LBD (n=70), AD (n=898), VBI (n=62), PPD (n=36) – that passed quality control and had an available scan matching a clinical visit <12 months apart. The PPD group is composed of subjects with different psychiatric disorders: depression (n=28), bipolar disorder (n=2), anxiety disorder (n=2) and other psychiatric diseases (n=5). Of note, subjects that did not have any cognitive and/or behavioral impairment (n=51) were excluded from this samples according to the following NACC variables: 1) NACCCOGF= 0 (No impairment in cognition), 2) COGMODE= 0 (No impairment in cognition), 3) NACCBEHF= 0 (No behavioral symptoms, for the bvFTD cohort), and 4) COMPORT= 0.0 (No impairment in behavior, comportment, and personality, for the bvFTD cohort).

**Figure 1.**
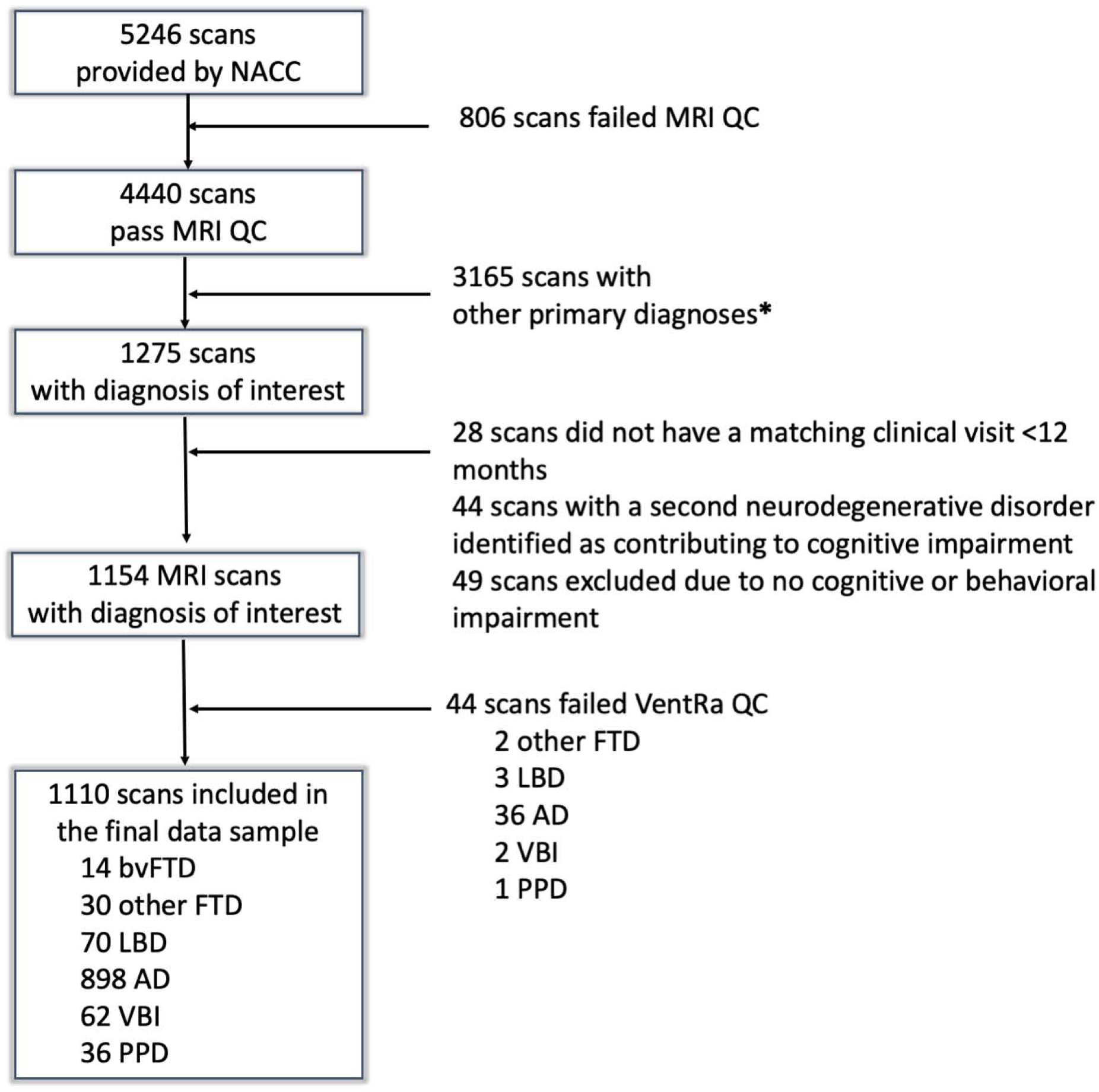
Flowchart showing the subject selection procedure. Abbreviations: bvFTD: behavioral variant frontotemporal dementia; FTD: Frontotemporal dementia; LBD: Lewy Body Disease; AD: Alzheimer’s dementia; VBI: Vascular Brain Injury; PPD: Primary Psychiatric Disorder. * 144 scans were excluded due to missing diagnosis, 1931 due to no cognitive impairment and, 1090 due to primary diagnoses other than the groups of interest (i.e., progressive supranuclear palsy, corticobasal degeneration, prion disease, traumatic brain injury, normal-pressure hydrocephalus, central nervous system neoplasm and other neurologic, genetic, or infectious condition).

### 2.2 MRI acquisition parameters

Out of all the participants selected for the final data sample, there were 529 3.0T and 449 1.5T scans acquired by either GE (N = 575), SIEMENS (N = 189) or Philips (N = 81) scanners (no data on the field strength was available for 132 scans). The scans were acquired in sagittal, coronal, and axial planes from either T1-weighted (T1w) Turbo Field Echo (TFE), Fast spin echo (FSE), fast spoiled gradient-echo (FSPGR) or Magnetization Prepared - RApid Gradient Echo (MPRAGE) sequences. Other acquisition parameters were echo time range: 0.0013-0.02; repetition time range: 0.006-3.00; number of slices in x range: 124-512; number of slices in y range: 54-512; and number of slices in z range: 29-512; slice thickness range: 0.8 – 5; voxel size x range: 0.4-1.2; voxel size y range: 0.4-3; and voxel size z range: 0.5-7.

### 2.3 Statistical analysis

All statistical analyses were conducted using MATLAB (version R2021b). Differences in categorical variables between the cohorts were assessed using chi-square and continuous variables were assessed using one-way ANOVA or Kruskal-Wallis variance analysis depending on the distribution of the variables based on a normality test. Post-hoc two-sample Student’s t-tests were conducted to examine clinical and ventricular differences. Results are expressed as mean ± standard deviation and median [interquartile range] as appropriate and *p* values for all the volumetric analyses have been corrected for multiple comparisons using the Bonferroni method so that any reported adjusted *p*-value < 0.05 is statistically significant.

### 2.4 VentRa tool

VentRa takes as input a comma separated (.csv) file providing the path for the raw T1-weighted images as well as age and sex of the subjects, and provides as output preprocessed images along with ventricle segmentations, QC files for the segmentations, as well as a .csv file including the diagnosis (based on the classifier trained on bvFTD vs the mixed group data) along with all the extracted ventricle features: i.e. total ventricle volume, ventricle volumes in each lobe and hemisphere, antero-posterior and left/right ratios. VentRa performs the classification task by means of a support vector machine classifier (fitcsvm function from MATLAB, with default parameters: linear kernel, Sequential Minimal Optimization) using ventricular features (APR, TVV, LRFR and LRTR) in combination with age and sex. This classifier was originally trained on a separate dataset^6^ and it thus tested here, unchanged, on an independent dataset.

#### 2.4.1 Ventricular feature estimation and analyses

VentRa includes a previously validated patch-based label fusion technique to segment the lateral ventricles^8^. The method uses expert manual segmentations as priors and estimates the label of each test subject voxel by comparing its surrounding 3D voxel patch against all the patches from the training library and performing a weighted label fusion using the intensity-based distances between the patch under study and the patches in the training subjects. All resulting segmentations were visually assessed by an experienced rater blind to the clinical diagnosis, and the incomplete/inaccurate segmentations (N=44 or 3.96%) were excluded. Using the Hammers brain atlas delineating frontal, parietal, temporal, and occipital lobes separately in the left and right hemispheres ^10 11^, lateral ventricular volumes were calculated per each brain lobe and hemisphere. Using coronal coordinate y=-12mm in the stereotaxic space (i.e., registered to the template) ventricles were divided into anterior and posterior portions to obtain the ventricular APR^6^. All volumes were normalized for intracranial volume and these ratios were log-transformed to achieve normal distribution.

## 3. RESULTS

### 3.1 Demographics

Table 1 shows the demographic and cognitive testing performances for all the cohorts. The bvFTD subjects were younger than LBD, AD and VBI subjects, and the age difference was not significant between bvFTD, other FTD and PPD. There were no significant differences in years of cognitive impairment onset between the groups. There were no significant differences between bvFTD and all the other cohorts in Mini-Mental Status Examination (MMSE) (*p* >0.05). bvFTD subjects were slightly more severely impaired than AD according to Clinical Dementia Rating-Sum of Boxes (CDR-SB) (*p* = 0.05) and significantly more impaired than other FTD (*p*=0.01), VBI (*p*<0.001) and PPD (*p*<0.001). However, on average they remained mild cases (CDR 1.0) of bvFTD and at a stage relevant for differential diagnosis. Other cognitive scores can be found in Table 1.

**Table 1.** Demographic and clinical characteristics for all the cohorts. Values express Mean± SD / Median [interquartile range]. P value level of significance: 0.05. Abbreviations: bvFTD: behavioral variant frontotemporal dementia; FTD: Frontotemporal dementia; LBD: Lewy Body Disease; AD: Alzheimer’s dementia; VBI: Vascular Brain Injury; PPD: Primary Psychiatric Disorder; MMSE: Mini-Mental Status Examination; CDR: Clinical Dementia Rating; CDR-SB: Clinical Dementia Rating-Sum of Boxes. The bold values represent significant differences with the bvFTD cohort.

|  | bvFTD<br>(N=16) | Other FTD<br>(N=30) | LBD<br>(N=70) | AD<br>(N=898) | VBI<br>(N=62) | PPD<br>(N=36) | *p*<br>value |
| --- | --- | --- | --- | --- | --- | --- | --- |
| *Age, y* | 62±7 | 66±10 | 74±7 | 75±9 | 79±8 | 69±10 | <0.001 |
| *Sex, male (%)* | 9 (56%) | 13 (43%) | 59 (84%) | 441 (49%) | 39 (63%) | 19 (53%) | <0.001 |
| *Years from cognitive impairment onset, y* | 2.8 [2.3-6.3] | 3 [2.3-5.5] | 4.6 [3.3-6.8] | 4.3 [2.8-6.4] | 3.9 [2.1-6] | - | 0.08 |
| *MMSE (n= 860)* | 24 [21-28] | 25 [21-28] | 26 [20-28] | 24 [20-27] | 27 [24-28] | 28 [26-29] | <0.001 |
| *CDR Global score (n=1110)* | 1 [0.5-1] | **0.5 [0.5-1]** | 0.5 [0.5-1] | 0.5 [0.5-1] | **0.5 [0.5-1]** | **0.5 [0.5-0.5]** | <0.001 |
| *CDR-SB (n=1110)* | 5.25 [4.5-9] | **2.75 [1-4.5]** | 3 [2-7] | **3.5 [1.5-5]** | **1.5 [1-2.5]** | **1[0.5-2]** | <0.001 |
| *TRAIL-A time, s (n=1041)* | 37 [23-64] | 45 [33-63] | 56 [40-85] | 44 [32-66] | 55 [41-75] | 38 [31-54] | <0.001 |
| *TRAIL-B time, s (n=902)* | 76 [66-90] | 111 [81-179] | **171 [111-300]** | **149 [99 -266]** | **204 [116-292]** | 103 [64-170] | <0.001 |
| *Digits forward correct trials (n=852)* | 6 [5 -9] | 6 [4-6] | 8 [7-9] | 7 [6-9] | 7 [6-8] | 8 [6-10] | 0.01 |
| *Digits backwards (n=850)* | 5 [4-7] | 5 [4-6] | 4 [3-5] | 5 [4-6] | 5 [4-6] | 6 [4-7] | 0.04 |
| *WAIS (n=767)* | 43 [29-46] | 38 [24-48] | 26 [21-35] | 32 [22-42] | 29 [19-34] | 35 [27-45] | 0.002 |
| *BOSTON (n=443)* | 20 [17-23] | 22 [11-29] | 24 [21-26] | 22 [16-25] | 23 [18-26] | 26 [21-27] | 0.02 |
| *Semantic Fluency – Animals (n=1081)* | 11 [4-13] | 11 [6-18] | 12 [7-16] | 12 [9-16] | 13 [11-16] | **16 [13-19]** | <0.001 |
| *Semantic Fluency – Vegetables (n=1078)* | 6 [3-8] | 8 [4-12] | 7 [1-10] | 8 [5-11] | 10 [7-11] | **12 [9-16]** | <0.001 |

### 3.2 Classification task with VentRa

#### 3.2.1 bvFTD vs PPD

VentRa achieved an accuracy of 84%, 71% sensitivity and 89% specificity in discriminating bvFTD subjects from PPD based on ventricular features (Figure 2). Positive and negative likelihood ratios (LR) were 6.43 and 0.31 respectively.

**Figure 2.**
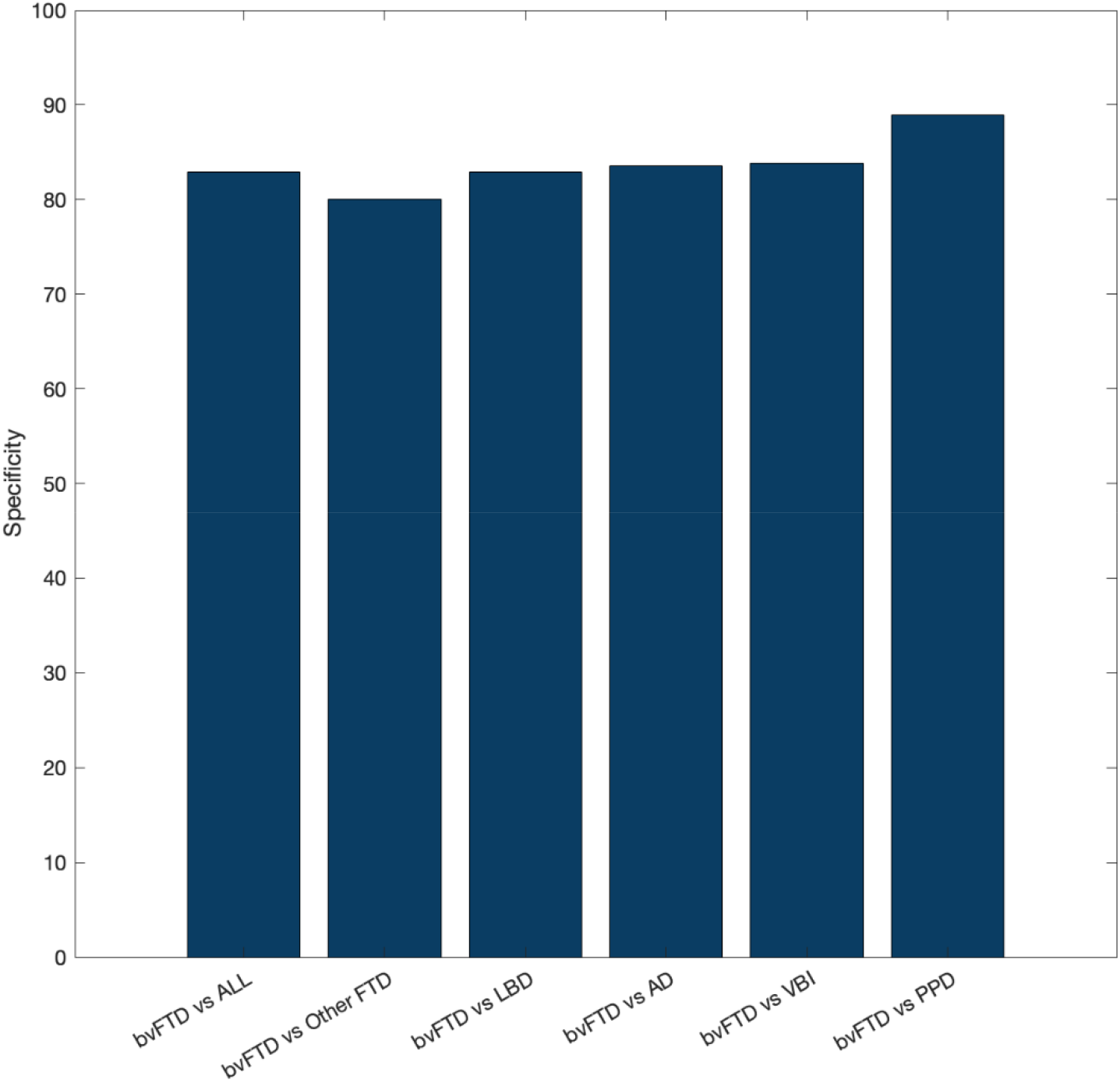
Specificity for the different classification tasks. Abbreviations: bvFTD: behavioral variant frontotemporal dementia; FTD: Frontotemporal dementia; LBD: Lewy Body Disease; AD: Alzheimer’s dementia; VBI: Vascular Brain Injury; PPD: Primary Psychiatric Disorder

The classification using VentRa resulted in 29% rate of false negatives (FN). As shown in Figure 3, these subjects were older than the bvFTD subjects correctly classified (63±8 vs 72±3 years respectively, p=0.06) and they had significantly smaller ventricular APR (*p*=0.01). The FN also performed significantly better than the true positives (TP) on MMSE (*p*=0.02). Yet, the disease duration was not significantly different than true positives (*p*=0.2) nor were the severity according to CDR global score (Median CDR Global_TP=_ 1 and Median CDR Global_FN=_ 0.75, *p*=0.2) and CDR-SB (Median CDR-SB_TP_= 6.5 and Median CDR-SB_FN_=5, *p*=0.2). Finally, no significant differences were found in severity of behavior, comportment, and personality impairment (*p*=0.4) according to NACC variable ‘COMPORT’ where 0= No impairment; 0.5= Questionable impairment; 1= Mild impairment; 1.5= Moderate impairment; and 2= Severe impairment).

**Figure 3.**
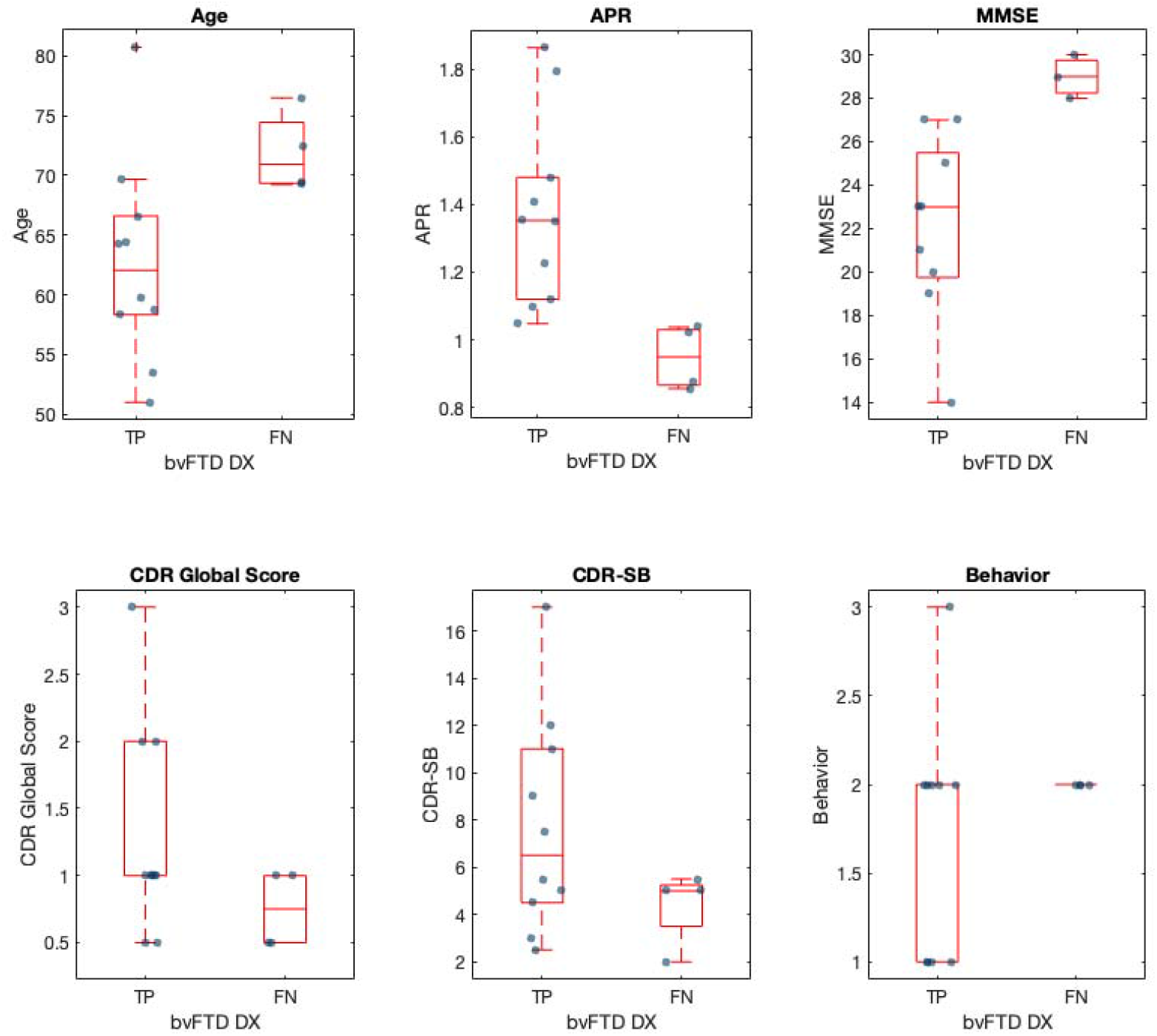
Boxplots showing the differences between bvFTD subjects correctly classified with VentRa (True Positives) and those classified as not compatible with bvFTD (False Negatives). Abbreviations: bvFTD: behavioral variant frontotemporal dementia; TP= True Positives; FN: False Negatives; APR: anteroposterior ratio; MMSE: Mini-Mental Status Examination; CDR: Clinical Dementia Rating; CDR-SB: Clinical Dementia Rating-Sum of Boxes. Behavior refers to a NACC variable ‘COMPORT (behavior, comportment, and personality impairment) where 0= No impairment; 0.5= Questionable impairment; 1= Mild impairment; 1.5= Moderate impairment; and 2= Severe impairment.

On the other hand, the false positive (FP) rate was 11%. When comparing the FP against the PPD subjects correctly diagnosed as not compatible with bvFTD (Figure 4), no significant differences were found in age (*p*=0.9), MMSE (*p*=0.8), CDR (*p*=0.4), CDR-SB (*p*=0.2) or severity of behavioral impairment (‘COMPORT’, *p*=0.33). However, the FP showed statistically significant larger APR compared to subjects correctly classified (1.26±0.3 and 0.97±0.3 respectively, *p*=0.04).

**Figure 4.**
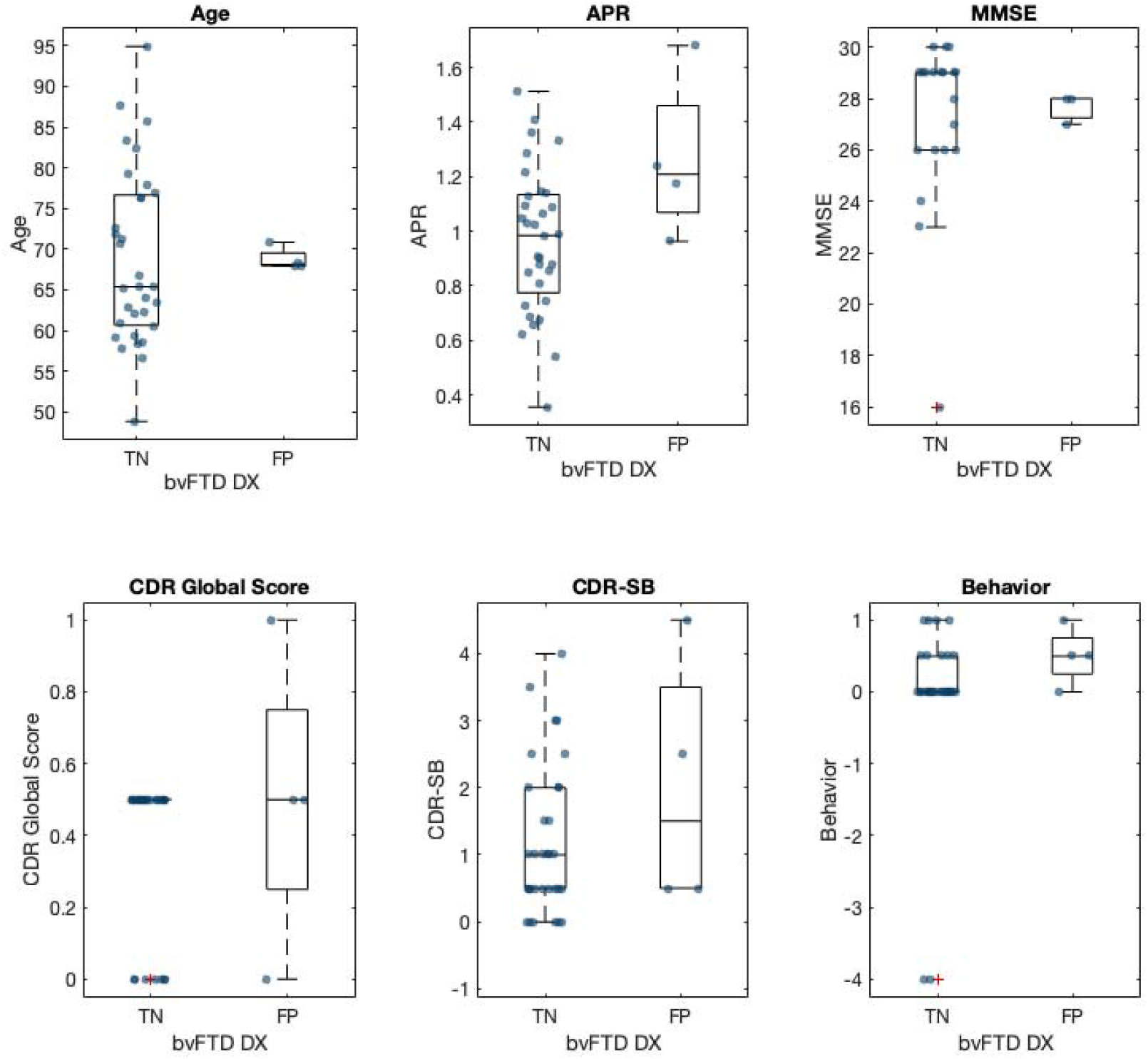
Boxplots showing the differences between PPD subjects incorrectly classified as bvFTD with VentRa (False Positives) and those classified as not compatible with bvFTD (True Negatives). Abbreviations: bvFTD: behavioral variant frontotemporal dementia; PPD: Primary psychiatric disorder; TN= True Negatives; FN: False Positives; APR: anteroposterior ratio; MMSE: Mini-Mental Status Examination; CDR: Clinical Dementia Rating; CDR-SB: Clinical Dementia Rating-Sum of Boxes. Behavior refers to a NACC variable ‘COMPORT (behavior, comportment, and personality impairment) where 0= No impairment; 0.5= Questionable impairment; 1= Mild impairment; 1.5= Moderate impairment; and 2= Severe impairment.

#### 3.2.2 bvFTD vs other neurodegenerative cohorts

Using ventricular features to identify bvFTD from a mixed age-matched cohort (‘ALL’= Other FTD, LBD, AD, VBI and PPD) VentRa was able to correctly classify other disorders as ‘not compatible with bvFTD’ with a specificity of 83%. The specificity against each of the other individual cohorts were 80% for other FTD, 83% for LBD, 83% for AD and 84% for VBI (Figure 2)

## 4. DISCUSSION

In the present study, we aimed to validate our previously developed automated tool VentRa^6^ for the estimation of ventricular features on T1w MRI for the individual prediction of bvFTD within a mixed cohort of subjects from an independent dataset of clinically acquired MRIs. The main findings were: 1) VentRa was able to accurately differentiate bvFTD from PPD (84% accuracy, 71% sensitivity, 89% specificity); 2)VentRa was able to identify bvFTD from a mixed cohort with a specificity of 83%; 3) From each of the other individual cohorts (other FTD, LBD, AD and VBI), VentrRa correctly identified between 80-84% as not compatible with bvFTD; 4) Our results show good positive and negative likelihood ratios proving its reliability to rule in and out bvFTD among the wider group of patients exhibiting behavioral changes such as apathy, disinhibition, and loss of empathy^12^.

While current clinical diagnostic criteria for the different neurodegenerative disorders in combination with several available pathologic biomarkers (i.e. AD biomarkers in blood/cerebrospinal fluid and Positron emission tomography) perform reasonably well, the main clinical challenge in the differential diagnosis of bvFTD is still represented by PPD. Clinical criteria for bvFTD are less helpful to distinguish it from PPD^2 13^. No bedside cognitive screening has yet been able to clearly discriminate bvFTD from PPD. Neuropsychological assessment frequently focuses more on the executive dysfunction characteristic of bvFTD though it is highly nonspecific for FTD versus PPDs^13^. Despite suggestions of its efficacy, dedicated assessment of social cognition is often not included in standard neuropsychological assessment batteries^13 14^. Furthermore, several social cognitive instruments that have been developed for research purposes are yet to have normative data available^15^.

Neuroimaging is already crucial for bvFTD diagnosis; frontal and/or anterior temporal atrophy or hypometabolism increases certainty from possible to probable bvFTD in current criteria^16^. Yet, MRIs may not be obviously abnormal in early stages or in certain variants of bvFTD, such as the C9orf72 phenotype^13 17^. In the Late Onset Frontal Lobe Syndrome (LOF) study, the standard visual review of structural MRI was found to be useful for the differential diagnosis of bvFTD versus PPD, however only had 70% sensitivity ^18 19^. It is hypothesized that volumetric MRI could improve this sensitivity, however there is insufficient evidence currently to formally recommend volumetric analytic techniques in clinical populations with behavioral changes^15^. Furthermore, while some machine learning algorithms using MRI volumetry perform well against controls and AD cohorts, less is known about the performance against PPD. Zhutovsky et. al. performed a classification of bvFTD vs PPD with different data usage approaches including clinical, MRI voxel-wise, MRI-Region of Interest (ROI), Clinical + MRI ROI, and Clinical + MRI voxel-wise techniques^20^. In their study, a support vector machine was trained on a sample of 73 patients (18 bvFTD, 28 neurological and 27 psychiatric). Zhutovsky et. al. report cross-validation accuracies of 82.1% (76.7% sensitivity, 87.5 specificity) and 78.3% (75.9% sensitivity, 80.8% specificity) using only MRI voxel-wise and MRI ROI approaches respectively. Cross-validation can overestimate performance as the same cohort is used to select features and optimize the method’s hyperparameters. In contrast, our validation using *independent* data, our method generalizes well as we achieved 84% accuracy, 71% sensitivity, 89% specificity in discriminating bvFTD subjects from PPD with VentRa. This accuracy with higher specificity relative to sensitivity is in line with reported accuracy from expert neuroradiological visual review reported in the LOF cohort^18^. Although the performance of VentRa was not superior, the fact that it is completely automated and performs as well as academic centre experts could improve clinical practice particularly in community areas. Further, VentRa has the potential to improve over time by including more training data.

Still, our results show a FN rate of 29%. This is likely because these FN bvFTD cases are older than the TP cases and outside of the age range of bvFTD subjects from the Frontotemporal Lobar Degeneration Neuroimaging Initiative (FTLDNI) that were originally used to train VentRa (Age ranges: bvFTD_NIFD_ 61±6 y, TP_NACC_ 63±8 y, and FN_NACC_ 72±3 y). Since these subjects were outside the operating range of the classifier, they were therefore more likely to be misclassified, thus underlying the importance of having training data that well represents the population of interest. It is highly probable that adding extended ages to the training dataset will likely improve the results. Not only were the FN older, but also they had better cognition (MMSE, CDR Global and CDR-SB) and their ventricular APR was smaller, we might therefore argue that there could be a mismatch between the clinical population included in NACC (including some older and milder cases, possibly with more ambiguous diagnoses) and the FTLDNI data (more advanced and clear cut cases) used to train VentRa. We also need to consider the fact that in older age FTD there is a higher prevalence of mixed pathologic diagnoses^21^, and therefore a classification algorithm is unlikely perform as well in this population.

Our study has two main strengths. First, VentrRa has been trained and cross-validated on a sample composed from two different multi-center databases (FTLDNI^22^ and Alzheimer’s Disease Neuroimaging Initiative-ANDI-^23^) and its performance was tested here on a held-out database which also included multi-center and multi-scanner data from different scanner models of both 1.5T and 3T field strengths (NACC). In addition, the failure rate of the VentRa QC was very low (4%), particularly considering the variability of the MRI acquisition parameters (i.e., slice thickness range: 0.8 – 5; voxel size x range: 0.4-1.2; voxel size y range: 0.4-3; and voxel size z range: 0.5-7). This further reinforces the generalizability (i.e., external validity) of our results and ensures their applicability in a clinical scenario with different scanners and with different magnetic field strengths. This raises the confidence that VentRa would work reliably in clinical settings. Secondly, our tool is based on standard structural T1w MRI and uses a combination of easily measurable features of the lateral ventricles which can be reliably segmented using a variety of publicly available tools such as FreeSurfer in addition to the patch-based method used in VentRa^7 8^. Further, all the image processing tools used in this study have been established and validated for use in multi-center and multi-scanner datasets ^11 24-28^, and have been designed to minimize such center- and scanner-related differences. The ventricular features are much less likely to be impacted by such differences, particularly for the APR, where any such discrepancies would be minimized and perhaps cancelled in the ratio.

Our study is not without limitations. The number of bvFTD subjects with available and good quality scans was relatively limited. In the future, it will be important to evaluate the robustness and generalizability of our results in other bvFTD cohorts. In addition, it will be valuable to see how well VentRa would perform against a more heterogenous PPD cohort and to predict future course in patients with clinically ambiguous FTD presentations (i.e., predict who has a neurodegenerative underlying pathology versus who has PPD). Since we have made both the image processing pipeline and classification model publicly available with VentRa (http://nist.mni.mcgill.ca/VentRa/), this can be easily achieved by others who might have access to such datasets.

To conclude, these results prove the robustness and generalizability of VentRa, indicating the potential usefulness of this automated MRI-based tool for improving the diagnostic certainty of bvFTD. If validated in a prospective larger sample, VentRa has the potential to improve diagnostic accuracy, particularly against populations with behavioral and/or psychiatric symptoms enabling reduced diagnostic and intervention time, particularly in settings with less access to specialized neuroradiology.

## Data Availability

All data produced in the present study are available upon reasonable request to the authors. Complete NACC dataset can be obtained upon request (https://naccdata.org/requesting-data/data-request-process)

## Acknowledgements

MD reports receiving research funding from the Healthy Brains for Healthy Lives (HBHL), Alzheimer Society Research Program (ASRP), and Douglas Research Centre (DRC).

DLC reports receiving research funding from Canadian Institutes of Health research, the Canadian National Science and Engineering Research Council, Brain Canada, the Weston Foundation, and the Famille Louise & André Charron.

SD receives salary funding from the Fonds de Recherche du Québec – Santé (FRQS)

The NACC database is funded by NIA/NIH Grant U24 AG072122. NACC data are contributed by the NIA-funded ADCs: P50 AG005131 (PI James Brewer, MD, PhD), P50 AG005133 (PI Oscar Lopez, MD), P50 AG005134 (PI Bradley Hyman, MD, PhD), P50 AG005136 (PI Thomas Grabowski, MD), P50 AG005138 (PI Mary Sano, PhD), P50 AG005142 (PI Helena Chui, MD), P50 AG005146 (PI Marilyn Albert, PhD), P50 AG005681 (PI John Morris, MD), P30 AG008017 (PI Jeffrey Kaye, MD), P30 AG008051 (PI Thomas Wisniewski, MD), P50 AG008702 (PI Scott Small, MD), P30 AG010124 (PI John Trojanowski, MD, PhD), P30 AG010129 (PI Charles DeCarli, MD), P30 AG010133 (PI Andrew Saykin, PsyD), P30 AG010161 (PI David Bennett, MD), P30 AG012300 (PI Roger Rosenberg, MD), P30 AG013846 (PI Neil Kowall, MD), P30 AG013854 (PI Robert Vassar, PhD), P50 AG016573 (PI Frank LaFerla, PhD), P50 AG016574 (PI Ronald Petersen, MD, PhD), P30 AG019610 (PI Eric Reiman, MD), P50 AG023501 (PI Bruce Miller, MD), P50 AG025688 (PI Allan Levey, MD, PhD), P30 AG028383 (PI Linda Van Eldik, PhD), P50 AG033514 (PI Sanjay Asthana, MD, FRCP), P30 AG035982 (PI Russell Swerdlow, MD), P50 AG047266 (PI Todd Golde, MD, PhD), P50 AG047270 (PI Stephen Strittmatter, MD, PhD), P50 AG047366 (PI Victor Henderson, MD, MS), P30 AG049638 (PI Suzanne Craft, PhD), P30 AG053760 (PI Henry Paulson, MD, PhD), P30 AG066546 (PI Sudha Seshadri, MD), P20 AG068024 (PI Erik Roberson, MD, PhD), P20 AG068053 (PI Marwan Sabbagh, MD), P20 AG068077 (PI Gary Rosenberg, MD), P20 AG068082 (PI Angela Jefferson, PhD), P30 AG072958 (PI Heather Whitson, MD), P30 AG072959 (PI James Leverenz, MD)

## ABBREVIATIONS

bvFTD: Behavioral Variant Frontotemporal Dementia
PPD: Primary Psychiatric Disorder
DBM: deformation-based morphometry
CN: normal controls
APR: anteroposterior ratio
MCI: mild cognitive impairment
AD: Alzheimer’s dementia
FTD: Frontotemporal Dementia
TVV: total ventricular volume
LRFR: left-right frontal ratio
LRTR: left-right temporal ratio
NACC: National Alzheimer’s Coordinating Center dataset
LBD: Lewy Body Disease
VBI: Vascular Brain Injury
T1w: T1-weighted (T1w)
TFE: Turbo Field Echo
FSE: Fast spin echo
FSPGR: fast spoiled gradient-echo
MPRAGE: Magnetization Prepared - RApid Gradient Echo
MMSE: Mini-Mental Status Examination
CDR: Clinical Dementia Rating
CDR-SB: Clinical Dementia Rating-Sum of Boxes
LR: Likelihood ratios
FN: False negatives
TP: True positives
FP: False positives
ROI: Region of Interest

